# Clinical Mortality Review in a Large COVID-19 Cohort

**DOI:** 10.1101/2020.08.05.20168146

**Authors:** Mark P. Jarrett, Susanne E. Schultz, Julie S. Lyall, Jason J. Wang, Lori Stier, Marcella De Geronimo, Karen L. Nelson

**Affiliations:** Donald and Barbara Zucker School of Medicine at Hofstra/Northwell, Hempstead, New York; Institute for Clinical Excellence and Quality/Safety, Northwell Health, New Hyde Park, New York; Krasnoff Quality Management Institute, Northwell Health, New Hyde Park, New York

## Abstract

**Background:** Northwell Health (Northwell), an integrated health system in New York, treated more than 15000 inpatients with coronavirus disease (COVID-19) at the US epicenter of the severe acute respiratory syndrome coronavirus 2 (SARS-CoV-2) pandemic. We describe the demographic characteristics of COVID-19 mortalities, observation of frequent rapid response teams (RRT)/cardiac arrest (CA) calls for non-intensive care unit (ICU) patients, and factors that contributed to RRT/CA calls.

**Methods:** A team of registered nurses reviewed medical records of inpatients who tested positive for SARS-CoV-2 via polymerase chain reaction (PCR) before or on admission and died between March 13 (first Northwell inpatient expiration) and April 30, 2020 at 15 Northwell hospitals. Findings for these patients were abstracted into a database and statistically analyzed.

**Findings:** Of 2634 COVID-19 mortalities, 56.1% had oxygen saturation levels greater than or equal to 90% on presentation and required no respiratory support. At least one RRT/CA was called on 42.2% of patients at a non-ICU level of care. Before the RRT/CA call, the most recent oxygen saturation levels for 76.6% of non-ICU patients were at least 90%. At the time RRT/CA was called, 43.1% had an oxygen saturation less than 80%.

**Interpretation:** This study represents one of the largest cohorts of reviewed mortalities that also captures data in non-structured fields. Approximately 50% of deaths occurred at a non-ICU level of care, despite admission to the appropriate care setting with normal staffing. The data imply a sudden, unexpected deterioration in respiratory status requiring RRT/CA in a large number of non-ICU patients. Patients admitted to a non-ICU level of care suffer rapid clinical deterioration, often with a sudden decrease in oxygen saturation. These patients could benefit from additional monitoring (eg, continuous central oxygenation saturation), although this approach warrants further study.

**Funding:** National Institute on Aging and the National Library of Medicine of the National Institutes of Health.

**RESEARCH IN CONTEXT:** *Evidence before the study:* The world first learned of SARS-CoV-2 through a landmark study published by the Lancet in January 2020. Early evidence was limited due to the novel nature of the virus, clinician inexperience in treating COVID-19, small sample size of primarily hospital inpatients in early observational studies, use of structured datasets for data collection, and early and evolving treatment guidelines. We collected evidence from leading medical journals from January 2020 through June 2020 that primarily describe the clinical characteristics of COVID-19 (eg, patient age, sex, and comorbidities) as well as the different approaches to treatment that were being used. Our search consisted of real-time publications as they became available. Analyzing the characteristics of patients who died can help to define the clinical nature of COVID-19 and potentially suggest new care protocols.

*Added value of this study:* Unlike prior research, our study represents one of the largest cohorts of COVID-19 mortalities abstracted from both structured and unstructured fields in the medical record using data collected during the surge of infections in the New York metropolitan area in March through April 2020. Our study identified an unusual pattern of respiratory decompensation in patients who presented to the hospital with acceptable oxygen saturation levels and were therefore admitted to a non-ICU setting for care. Through our unique analysis, we identified a cohort of patients who experienced a rapid response team/cardiac arrest call after a sudden and unexpected decrease in their oxygenation saturation levels.

*Implications of available evidence:* Our findings, based on over 2600 inpatient mortalities, suggest that there is a subpopulation of patients who are admitted to a non-ICU setting where continuous oxygenation saturation monitoring is not a standard, but may be warranted. Further research is needed to understand which patients are at highest risk for mortality, due to age and comorbidities, and the implications of continuous monitoring on mortality rates. This finding is relevant to a wide audience of national and international health care providers.

## INTRODUCTION

Downstate New York was the first epicenter of the severe acute respiratory syndrome coronavirus 2 (SARS-CoV-2) pandemic in the United States.^1-2^ Northwell Health (Northwell), an integrated health system, treated more than 15000 inpatients with coronavirus disease (COVID-19). Comprehensively analyzing the characteristics of those who died can help define the clinical nature of COVID-19 infection and potentially suggest new care protocols. For 7 years, Northwell has used a centralized mortality review process with data validated through rigorous internal review and inter-rater reliability (92% to 96%). This robust process used a customized database to review all 2634 COVID-19 mortalities in Northwell’s adult acute care hospitals between March and April 2020. During this overwhelming surge, documentation was in various notes, as well as in structured fields in the electronic health record (EHR) systems. This study describes the demographic characteristics of COVID-19 mortalities and the observation of frequent calls for rapid response teams (RRT)/cardiac arrest (CA) in patients not in the intensive care unit (ICU). It also discusses factors that contributed to the RRT/CA calls, which may be a significant element in planning for a pandemic resurgence.

## METHODS

Northwell is New York State’s largest health care provider and private employer. With 23 hospitals (including specialty hospitals) and nearly 800 outpatient practice sites, the organization cares for over 2 million people in greater metropolitan New York. A team of registered nurses in the corporate quality department retrospectively reviewed medical records from 15 acute care hospitals. This team routinely conducts clinical reviews of all adult acute inpatient mortalities-approximately 5000 per year. A physician advisor was available to the team to consult on clinical questions.

Database elements were based on Northwell’s experience with treating COVID-19 patients, literature review from countries that had early experience in treating patients, and clinical trials being conducted at the Feinstein Institutes for Medical Research. Also, the data were captured in the database established under the direction of critical care intensivists at the epicenter of the pandemic, other subject matter experts, and quality leadership. During data abstraction, modifications and enhancements were made to the database based on trends and emerging information. Demographics, comorbidities, clinical findings, and management of COVID-19 patients who died were analyzed.

Analyzed cases included inpatients who tested positive for SARS-CoV-2 via polymerase chain reaction (PCR) before or on admission and then died between March 13 (first Northwell inpatient death) and April 30, 2020. Emergency department (ED) mortalities were excluded. Demographic data and comorbidities were abstracted from the medical record on admitted patients. Initially data were collected on ten patient comorbidities that were deemed important and then narrowed down to six comorbidities for inclusion based on our initial analysis.

Transfers from one in-system hospital to another were merged and considered as a single visit. One patient outcome measured was a call for RRT/CA.

This Institutional Review Board of Northwell Health deemed this study as exempt and waived the requirement for informed consent.

Statistical analyses were performed using chi-square for categorical variables and *t*-test for continuous variables. A multivariable logistic regression model was created to determine independent risk factors for the outcome variable. Statistical significance was considered p<0.05. All statistical analyses were done in SAS v9.4 (SAS Institute).

## RESULTS

Baseline characteristics of 2634 COVID-19 mortalities are described in table 1. The age range was 21 to 107 years in the following categories: 21 to 39 (49; 1.86%), 40 to 59 (351; 13.3%), 60 to 79 (1241; 47.1%), and 80 and older (993; 37.7%). There were 1664 (63.2%) male and 970 (36.8%) female patients. Of all patients, 1256 (47.7%) were white, 463 (17.6%) black, 230 (8.7%) Asian, and 685 (26.0%) other/unknown. Insurance was Medicare for the majority of patients (1839; 69.8%). The most common comorbidities among those collected were hypertension (1719; 65.3%), diabetes mellitus (1043; 39.6%), and dementia (431; 16.4%). Fewer patients had chronic obstructive pulmonary disease (COPD) (385; 14.6%), heart failure (291; 11.1%), and end stage renal disease (166; 6.3%). Of these six comorbidities, more than 50% of patients had 2 or more comorbidities and 16.9% had 0 comorbidities. The majority of patients with a known body mass index (BMI) of 25 or more were categorized as follows: 25 to 29.99 (732; 27.8%), 30 to 34.99 (401; 15.2%), 35 to 39.99 (190; 7.2%), and 40 or higher (147; 5.6%). Most patients were admitted from home (1895; 71.9%). Of the remaining patients, 411 (15.6%) were admitted from a skilled nursing facility (SNF), 201 (7.6%) from an acute care facility, and 127 (4.8%) from rehabilitation. The number of patients with a prior ED visit within 7 days of admission was 125 (4.8%), and within 48 hours of admission was 51 (1.9%). The number of patients readmitted within 30 days was 194 (7.4%), within 7 days was 75 (2.9%), and within 24 hours was 20 (0.8%). On presentation, most (1478; 56.1%) patients had an oxygen saturation level greater than or equal to 90%, and 1397 (53.0%) required no respiratory support. Others required nasal cannula (363; 13.8%), non-rebreather (742; 28.2%), and mechanical ventilation (24; 0.9%). More than half of the patients who died (1403; 53.2%) required mechanical ventilation during their clinical course. Of those patients, 1332 (94.9%) had increasing oxygen requirements before intubation, 1259 (89.7%) were on traditional ventilators, 142 (10.1%) were on converted BiPAP machines, and 2 (0.1%) were on anesthesia machines. The length of time on mechanical ventilation was from 0 to 7 days for 851 (60.7%) patients and was 8 days or more for 552 (39.3%) patients.

**Table 1.**
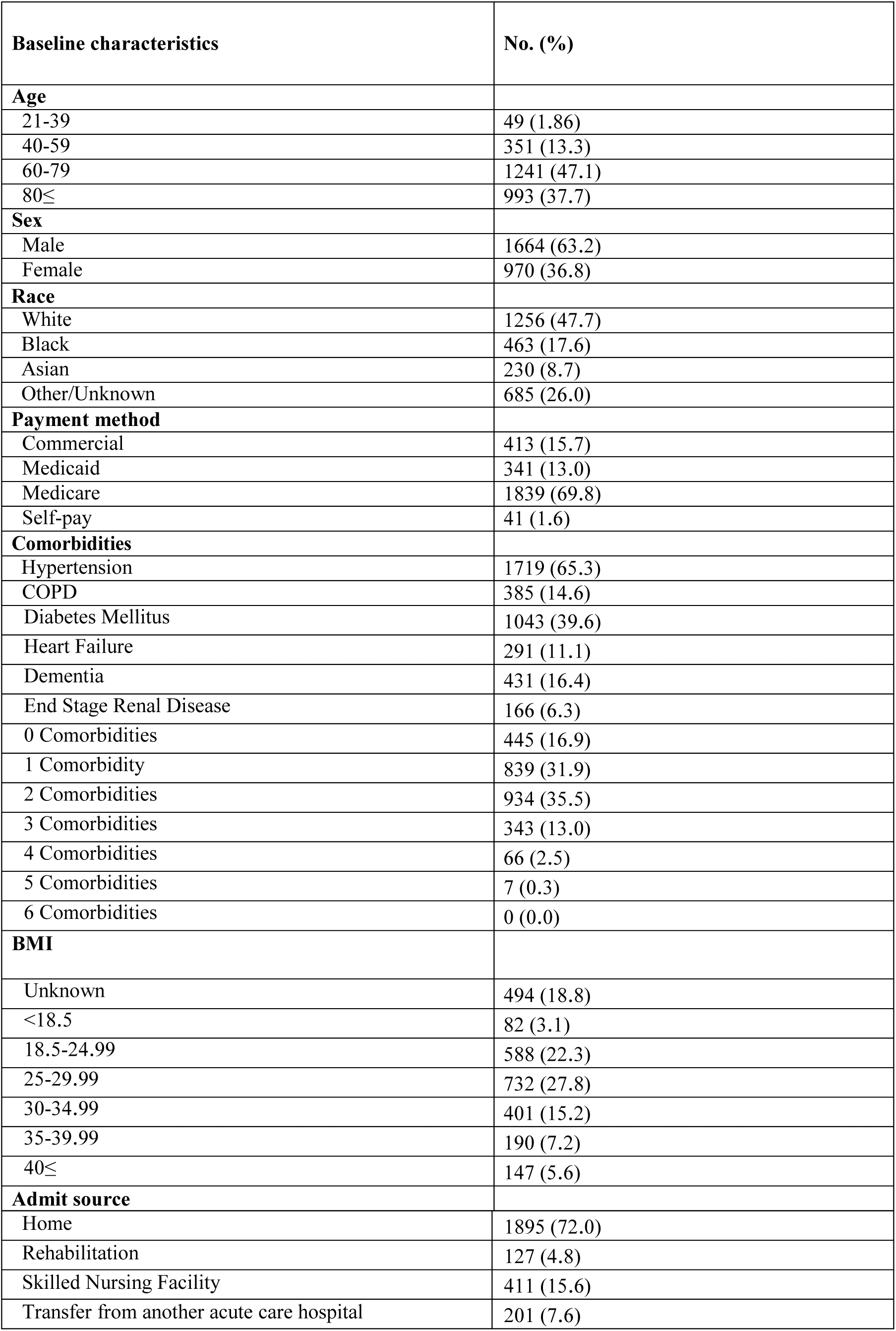
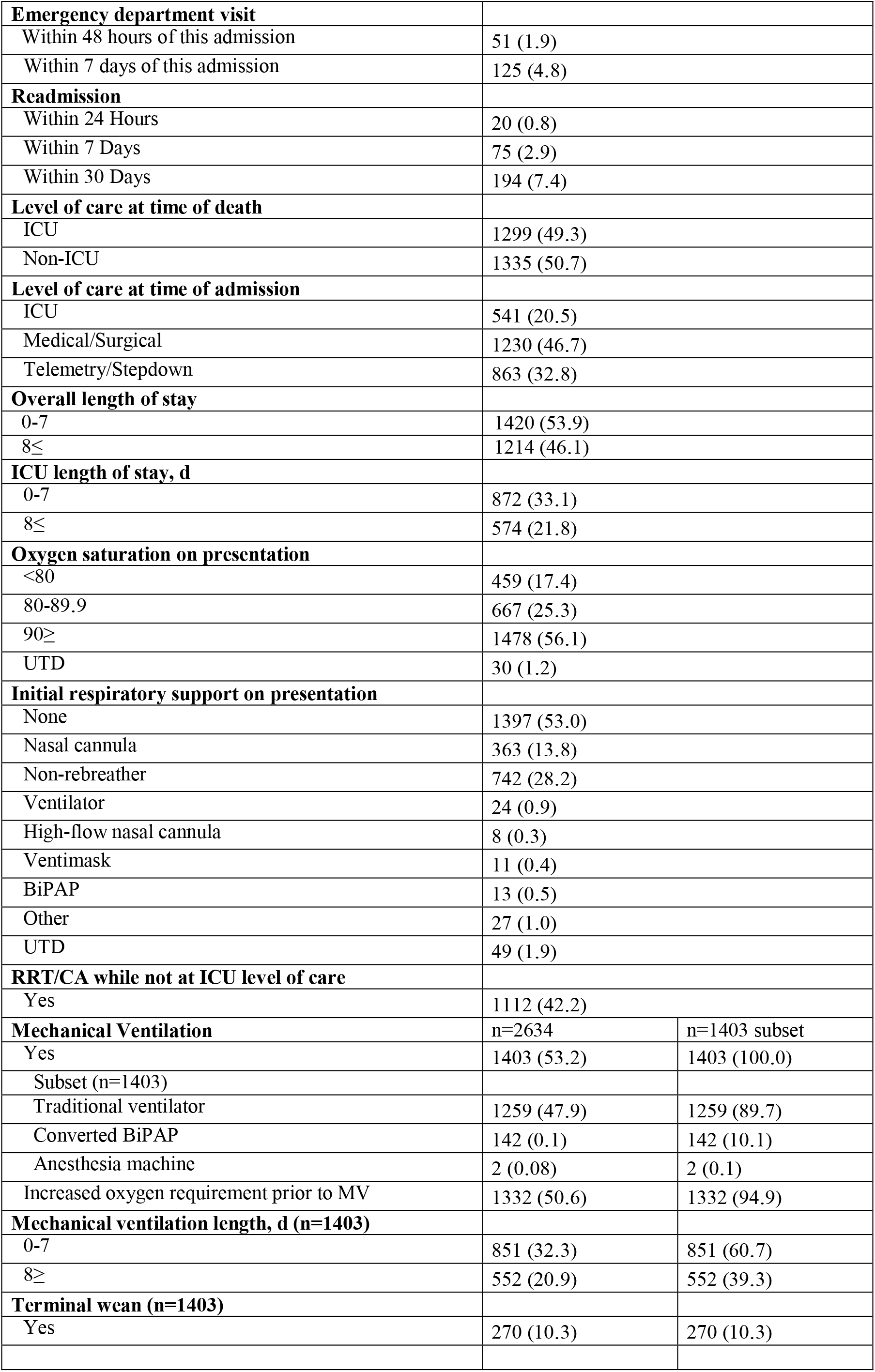
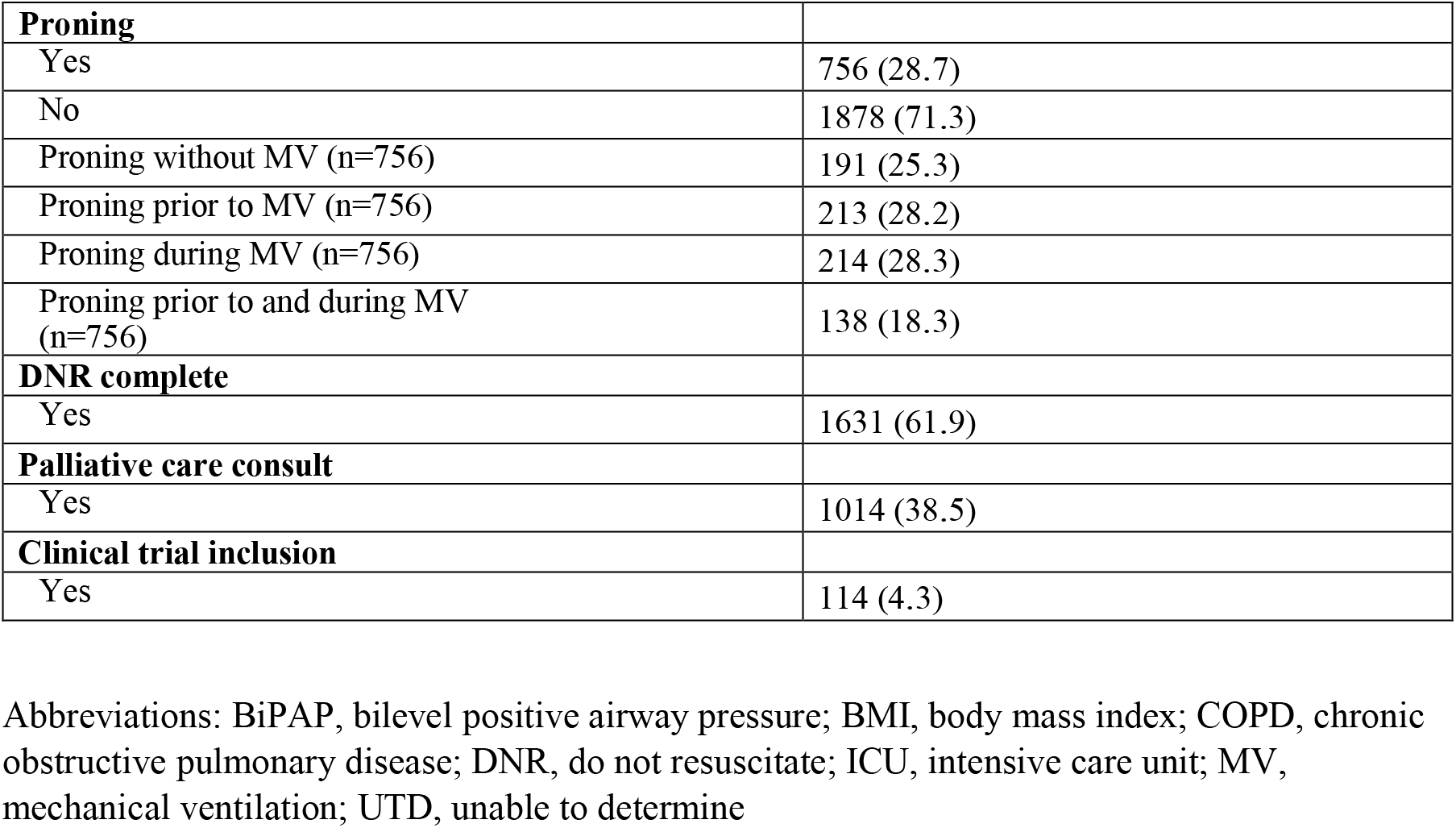
Baseline characteristics of Patients Hospitalized with COVID-19 Mortality (n=2634)

| **Baseline characteristics** | **No. (%)** |
| --- | --- |
| **Age** |  |
| 21-39 | 49 (1.86) |
| 40-59 | 351 (13.3) |
| 60-79 | 1241 (47.1) |
| 80≤ | 993 (37.7) |
| **Sex** |  |
| Male | 1664 (63.2) |
| Female | 970 (36.8) |
| **Race** |  |
| White | 1256 (47.7) |
| Black | 463 (17.6) |
| Asian | 230 (8.7) |
| Other/Unknown | 685 (26.0) |
| **Payment method** |  |
| Commercial | 413 (15.7) |
| Medicaid | 341 (13.0) |
| Medicare | 1839 (69.8) |
| Self-pay | 41 (1.6) |
| **Comorbidities** |  |
| Hypertension | 1719 (65.3) |
| COPD | 385 (14.6) |
| Diabetes Mellitus | 1043 (39.6) |
| Heart Failure | 291 (11.1) |
| Dementia | 431 (16.4) |
| End Stage Renal Disease | 166 (6.3) |
| 0 Comorbidities | 445 (16.9) |
| 1 Comorbidity | 839 (31.9) |
| 2 Comorbidities | 934 (35.5) |
| 3 Comorbidities | 343 (13.0) |
| 4 Comorbidities | 66 (2.5) |
| 5 Comorbidities | 7 (0.3) |
| 6 Comorbidities | 0 (0.0) |
| **BMI** |  |
| Unknown | 494 (18.8) |
| <18.5 | 82 (3.1) |
| 18.5-24.99 | 588 (22.3) |
| 25-29.99 | 732 (27.8) |
| 30-34.99 | 401 (15.2) |
| 35-39.99 | 190 (7.2) |
| 40≤ | 147 (5.6) |
| **Admit source** |  |
| Home | 1895 (72.0) |
| Rehabilitation | 127 (4.8) |
| Skilled Nursing Facility | 411 (15.6) |
| Transfer from another acute care hospital | 201 (7.6) |

|  |  |  |
| --- | --- | --- |
| **Emergency department visit** |  |  |
| Within 48 hours of this admission | 51 (1.9) |  |
| Within 7 days of this admission | 125 (4.8) |  |
| **Readmission** |  |  |
| Within 24 Hours | 20 (0.8) |  |
| Within 7 Days | 75 (2.9) |  |
| Within 30 Days | 194 (7.4) |  |
| **Level of care at time of death** |  |  |
| ICU | 1299 (49.3) |  |
| Non-ICU | 1335 (50.7) |  |
| **Level of care at time of admission** |  |  |
| ICU | 541 (20.5) |  |
| Medical/Surgical | 1230 (46.7) |  |
| Telemetry/Stepdown | 863 (32.8) |  |
| **Overall length of stay** |  |  |
| 0-7 | 1420 (53.9) |  |
| 8≤ | 1214 (46.1) |  |
| **ICU length of stay, d** |  |  |
| 0-7 | 872 (33.1) |  |
| 8≤ | 574 (21.8) |  |
| **Oxygen saturation on presentation** |  |  |
| <80 | 459 (17.4) |  |
| 80-89.9 | 667 (25.3) |  |
| 90≥ | 1478 (56.1) |  |
| UTD | 30 (1.2) |  |
| **Initial respiratory support on presentation** |  |  |
| None | 1397 (53.0) |  |
| Nasal cannula | 363 (13.8) |  |
| Non-rebreather | 742 (28.2) |  |
| Ventilator | 24 (0.9) |  |
| High-flow nasal cannula | 8 (0.3) |  |
| Ventimask | 11 (0.4) |  |
| BiPAP | 13 (0.5) |  |
| Other | 27 (1.0) |  |
| UTD | 49 (1.9) |  |
| **RRT/CA while not at ICU level of care** |  |  |
| Yes | 1112 (42.2) |  |
| **Mechanical Ventilation** | n=2634 | n=1403 subset |
| Yes | 1403 (53.2) | 1403 (100.0) |
| Subset (n=1403) |  |  |
| Traditional ventilator | 1259 (47.9) | 1259 (89.7) |
| Converted BiPAP | 142 (0.1) | 142 (10.1) |
| Anesthesia machine | 2 (0.08) | 2 (0.1) |
| Increased oxygen requirement prior to MV | 1332 (50.6) | 1332 (94.9) |
| **Mechanical ventilation length, d (n=1403)** |  |  |
| 0-7 | 851 (32.3) | 851 (60.7) |
| 8≥ | 552 (20.9) | 552 (39.3) |
| **Terminal wean (n=1403)** |  |  |
| Yes | 270 (10.3) | 270 (10.3) |
| **Proning** |  |
| Yes | 756 (28.7) |
| No | 1878 (71.3) |
| Proning without MV (n=756) | 191 (25.3) |
| Proning prior to MV (n=756) | 213 (28.2) |
| Proning during MV (n=756) | 214 (28.3) |
| Proning prior to and during MV (n=756) | 138 (18.3) |
| **DNR complete** |  |
| Yes | 1631 (61.9) |
| **Palliative care consult** |  |
| Yes | 1014 (38.5) |
| **Clinical trial inclusion** |  |
| Yes | 114 (4.3) |

Abbreviations: BiPAP, bilevel positive airway pressure; BMI, body mass index; COPD, chronic obstructive pulmonary disease; DNR, do not resuscitate; ICU, intensive care unit; MV, mechanical ventilation; UTD, unable to determine

Prone positioning was documented for 756 patients (28.7%), and 270 (10.3%) patients were terminally weaned. Do Not Resuscitate (DNR) orders were complete for 1631 (50.2%) patients. A palliative care consult was provided to 1014 (38.5%) patients. At the time of death, the level of care was the ICU for 1299 (49.3%) patients and non-ICU for 1335 (50.7%) patients.

A total of 1112 (42.2%) patients had a RRT/CA call at a non-ICU level of care versus 1522 (57.8%) who did not. As shown in table 2 and table 3, the RRT/CA group was significantly different from the non-RRT/CA group in terms of age, race, and comorbidities. There were 618 patients (55.6%) between 60 and 79 years old in the RRT/CA group compared to 623 patients (40.9%) in the non-RRT/CA group. In terms of race, white was significantly lower in the RRT/CA group (36.3% versus 55.9%; p<0.0001). The RRT/CA cohort had a significantly higher rate of diabetes mellitus (44.2% versus 36.3%, p<0.0001). Patients in the RRT/CA cohort were more likely to be admitted from home (926; 83.3%) versus the non-RRT/CA cohort (969; 63.7%). The RRT/CA cohort versus the non-RRT/CA cohort was more likely to be admitted to a medical/surgical unit [(576; 51.8%) versus (654; 42.9%)] or telemetry/step-down unit [(455; 40.9%) versus (408; 26.8%)], and to die at an ICU level of care [(671; 60.3%) versus (628; 41.3%)]. An overall length of stay (LOS) of 8 days or more was higher for the RRT/CA cohort (645; 58.0%) than the non-RRT/CA cohort (569; 37.4%), as was an ICU LOS of 0 to 7 days [(472; 42.0%) versus (400; 26.3%)] and 8 days or more [(271; 24.4%) versus (303; 19.9%)]. After adjusting for demographic and clinical characteristics, oxygen saturation levels at presentation were significant for the RRT/CA cohort at oxygen saturation levels of 80% to 89% [odds ratio (OR)=1.988; 95% CI: 1.511, 2.616] and of 90% or higher (OR=2.517; 95% CI: 1.962, 3.230). See logistic regression results (table 4).

**Table 2.** Baseline Characteristics of COVID-19 Mortalities that Experienced a RRT/CA at a Non-ICU Level of Care.

|  | **RRT/CA (n=2634)** |  |  |
| --- | --- | --- | --- |
| **Baseline characteristics** | **Yes, No. (%)**<br>**(n=1112)** | **No, No. (%)**<br>**(n=1522)** | **p value** |
| **Age** |  |  | <0.0001 |
| 21-39 | 19 (1.7) | 30 (2.0) |  |
| 40-59 | 194 (17.5) | 157 (10.3) |  |
| 60-79 | 618 (55.6) | 623 (40.9) |  |
| 80≤ | 281 (25.3) | 712 (40.8) |  |
| **Sex** |  |  | 0.3466 |
| Male | 714 (64.2) | 950 (62.4) |  |
| Female | 398 (35.8) | 572 (37.6) |  |
| **Race** |  |  | <0.0001 |
| White | 404 (36.3) | 852 (55.9) |  |
| Black | 235 (21.1) | 228 (15.0) |  |
| Asian | 125 (11.2) | 105 (6.9) |  |
| Other/Unknown | 348 (31.3) | 337 (22.1) |  |
| **Payment method** |  |  | <0.0001 |
| Commercial | 226 (20.3) | 187 (12.3) |  |
| Medicaid | 166 (14.9) | 175 (11.5) |  |
| Medicare | 702 (63.1) | 1137 (74.7) |  |
| Self-pay | 18 (1.6) | 23 (1.5) |  |
| **Comorbidities** |  |  |  |
| **Hypertension** |  |  | 0.2365 |
| Yes | 740 (66.5) | 979 (64.3) |  |
| No | 372 (33.5) | 543 (35.7) |  |
| **COPD** |  |  | 0.0828 |
| Yes | 147 (13.2) | 238 (15.6) |  |
| No | 965 (86.8) | 1284 (84.4) |  |
| Diabetes mellitus |  |  | <0.0001 |
| Yes | 491 (44.2) | 552 (36.3) |  |
| No | 621 (55.9) | 970 (63.7) |  |
| Heart failure |  |  | 0.0339 |
| Yes | 106 (9.5) | 185 (12.2) |  |
| No | 1006 (90.5) | 1337 (87.8) |  |
| Dementia |  |  | <0.0001 |
| Yes | 98 (8.8) | 333 (21.9) |  |
| No | 1014 (91.2) | 1189 (78.1) |  |
| End stage renal disease |  |  | 0.0154 |
| Yes | 85 (7.6) | 81 (5.3) |  |
| No | 1027 (92.4) | 1441 (94.7) |  |
| 0 Comorbidities | 202 (18.2) | 243 (15.9) | 0.4721 |
| 1 Comorbidity | 355 (31.9) | 484 (31.8) |  |
| 2 Comorbidities | 388 (34.9) | 546 (35.9) |  |
| 3 Comorbidities | 134 (12.1) | 209 (13.7) |  |
| 4 Comorbidities | 31 (2.8) | 35 (2.3) |  |
| 5 Comorbidities | 2 (0.2) | 5 (0.3) |  |
| **BMI** |  |  | <0.0001 |
| Unknown | 136 (12.2) | 358 (23.5) |  |
| <18.5 | 22 (1.9) | 60 (3.9) |  |
| 18.5-24.99 | 236 (21.2) | 352 (23.1) |  |
| 25-29.99 | 352 (31.7) | 380 (24.9) |  |
| 30-34.99 | 206 (18.5) | 195 (12.8) |  |
| 35-39.99 | 88 (7.9) | 102 (6.7) |  |
| 40≤ | 72 (6.5) | 75 (4.9) |  |
| **Admit source** |  |  | <0.0001 |
| Home | 926 (83.3) | 969 (63.7) |  |
| Rehabilitation | 34 (3.0) | 93 (6.1) |  |
| Skilled nursing facility | 80 (7.2) | 331 (21.7) |  |
| Transfer from another acute care Hospital | 72 (6.5) | 129 (8.5) |  |
| **Emergency department visit** |  |  |  |
| Within 48 hours of this admission |  |  | 0.0325 |
| Yes | 29 (2.6) | 22 (1.5) |  |
| No | 1083 (97.4) | 1500 (98.6) |  |
| Within 7 days of this admission |  |  | 0.1268 |
| Yes | 61 (5.5) | 64 (4.2) |  |
| No | 1051 (94.5) | 1458 (95.8) |  |
| **Readmission** |  |  |  |
| Within 24 hours |  |  | 0.5118 |
| Yes | 7 (0.6) | 13 (0.9) |  |
| No | 1105 (99.4) | 1509 (99.2) |  |
| Within 7 days |  |  | 0.8751 |
| Yes | 31 (2.8) | 44 (2.9) |  |
| No | 1081 (97.2) | 1478 (97.1) |  |
| Within 30 days |  |  | 0.0997 |
| Yes | 71 (6.4) | 123 (8.1) |  |
| No | 1041 (93.6) | 1399 (91.9) |  |
| **Level of care at time of death** |  |  |  |
| ICU | 671 (60.3) | 628 (41.3) |  |
| Non-ICU | 441 (39.7) | 894 (58.7) |  |
| **Level of care at time of admission** |  |  | <0.0001 |
| ICU | 81 (7.3) | 460 (30.2) |  |
| Medical/Surgical | 576 (51.8) | 654 (42.9) |  |
| Telemetry/Stepdown | 455 (40.9) | 408 (26.8) |  |
| **Overall length of stay, d** |  |  | <0.0001 |
| 0-7 | 467 (42.0) | 953 (62.6) |  |
| 8≤ | 645 (58.0) | 569 (37.4) |  |
| **ICU length of stay, d** |  |  | <0.0001 |
| 0-7 | 472 (42.5) | 400 (26.3) |  |
| 8≤ | 271 (24.4) | 303 (19.9) |  |
| **Oxygen saturation on presentation** |  |  | <0.0001 |
| <80 | 152 (13.7) | 307 (20.2) |  |
| 80-89.9 | 289 (26.0) | 378 (24.8) |  |
| 90≤ | 664 (59.7) | 814 (53.5) |  |
| UTD | 7 (0.6) | 23 (1.5) |  |
| **Initial respiratory support on presentation** |  |  | <0.0001 |
| None | 687 (61.8) | 710 (46.7) |  |
| Nasal cannula | 161 (14.5) | 202 (13.3) |  |
| High-flow nasal cannula | 0 (0.0) | 8 (0.5) |  |
| Ventimask | 2 (0.2) | 9 (0.6) |  |
| BiPAP | 2 (0.2) | 11 (0.7) |  |
| Non-rebreather | 239 (21.5) | 503 (33.1) |  |
| Ventilator | 1 (0.1) | 23 (1.5) |  |
| Other | 4 (0.4) | 23 (1.5) |  |
| UTD | 16 (1.4) | 33 (2.2) |  |
| **Mechanical ventilation** |  |  |  |
| Yes | 723 (65.0) | 680 (44.7) | <0.0001 |
| Traditional ventilator | 650 (58.5) | 609 (40.0) |  |
| Converted BiPAP | 71 (6.4) | 71 (4.7) |  |
| Anesthesia machine | 2 (0.2) | 0 (0.0) |  |
| Increased oxygen requirement before MV | 699 (62.9) | 633 (41.6) | <0.0001 |
| **Mechanical ventilation length, d** |  |  |  |
| 0-7 | 461 (41.5) | 390 (25.6) |  |
| 8≤ | 262 (23.6) | 290 (19.1) |  |
| **Terminal wean** |  |  | 0.5166 |
| Yes | 109 (9.8) | 161 (10.6) |  |
| No | 1003 (90.2) | 1361 (89.4) |  |
| **Proning** |  |  | <0.0001 |
| Yes | 500 (45.0) | 256 (16.8) |  |
| No | 612 (54.9) | 1266 (83.2) |  |
| Proning without MV | 116 (10.4) | 75 (4.9) |  |
| Proning before MV | 171 (15.4) | 42 (2.7) |  |
| Proning during MV | 99 (8.9) | 115 (7.5) |  |
| Proning before and during MV | 114 (10.3) | 24 (1.6) |  |
| **DNR complete** |  |  | <0.0001 |
| Yes | 558 (50.2) | 1073 (70.5) |  |
| No | 554 (49.8) | 449 (29.5) |  |
| **Palliative care consult** |  |  | 0.0005 |
| Yes | 385 (34.6) | 629 (41.3) |  |
| No | 727 (65.4) | 893 (58.7) |  |
| **Clinical trial inclusion** |  |  |  |
| Yes | 91(8.2) | 23(1.5) |  |
| No | 1021(91.8) | 1499 (98.5) |  |

Abbreviations: BiPAP, bilevel positive airway pressure; BMI, body mass index; COPD, chronic obstructive pulmonary disease; DNR, do not resuscitate; ICU, intensive care unit; MV, mechanical ventilation; RRT/CA, Rapid Response Team/Cardiac Arrest; UTD, unable to determine.

**Table 3.** Additional characteristics associated with RRT/CA While on a Non-ICU Level of Care (n=1112)

|  | **No. (%)** |
| --- | --- |
| **Required escalation in level of care following initial RRT/CA** |  |
| Yes | 716 (64.4) |
| **Oxygen saturation at time RRT/CA initiated** |  |
| <80 | 479 (43.1) |
| 80-89 | 407 (36.6) |
| 90≤ | 128 (11.5) |
| UTD | 98 (8.8) |
| **Oxygen supplement at time RRT/CA initiated** |  |
| Non-rebreather with or without nasal Cannula | 868 (78.1) |
| Nasal cannula | 147 (13.2) |
| Room air | 40 (3.6) |
| Venti mask | 18 (1.6) |
| Ventilator | 11 (1.0) |
| High-flow nasal cannula | 9 (0.8) |
| BiPAP | 5 (0.4) |
| UTD | 14 (1.3) |
| **Most recent oxygen saturation before RRT/CA initiated** |  |
| <80 | 43 (3.9) |
| 80-89 | 211 (18.9) |
| 90≤ | 852 (76.7) |
| UTD | 6 (0.5) |
| **Documented timing of most recent oxygen saturation before RRT/CA initiated, h** |  |
| <1 | 263 (23.7) |
| 1-2 | 191 (17.2) |
| 2-3 | 140 (12.6) |
| 3-4 | 109 (9.8) |
| 4< | 409 (36.8) |

Abbreviations: BiPAP, bilevel positive airway pressure; RRT/CA, Rapid Response Team/Cardiac Arrest; UTD, unable to determine.

**Table 4.** Regression Analysis of COVID-19 Mortalities that Experienced a RRT/CA at a Non-ICU Level of Care (n=2634)

| Baseline characteristics | Estimate | p value | Odds ratio | 95% CI Estimate |  |
| --- | --- | --- | --- | --- | --- |
| **Age** |  |  |  |  |  |
| 50-69 | 0.2653 | 0.1961 | 1.304 | 0.872 | 1.949 |
| 70-79 | 0.1721 | 0.4420 | 1.188 | 0.766 | 1.842 |
| 80≥ | -0.3179 | 0.1739 | 0.728 | 0.460 | 1.151 |
| **Sex** |  |  |  |  |  |
| Male | -0.2299 | 0.0170 | 0.795 | 0.658 | 0.960 |
| **Race** |  |  |  |  |  |
| Black | 0.6134 | <0.0001 | 1.847 | 1.445 | 2.361 |
| Asian | 0.6548 | <0.0001 | 1.925 | 1.395 | 2.655 |
| Other/Unknown | 0.5333 | <0.0001 | 1.704 | 1.362 | 2.133 |
| **Payment method** |  |  |  |  |  |
| Medicaid | -0.0458 | 0.7818 | 0.955 | 0.691 | 1.321 |
| Medicare | -0.0107 | 0.9397 | 0.989 | 0.750 | 1.305 |
| Self-pay | -0.3020 | 0.3972 | 0.739 | 0.367 | 1.488 |
| **Comorbidities** |  |  |  |  |  |
| Heart failure | 0.1429 | 0.3397 | 1.154 | 0.860 | 1.547 |
| End stage renal Disease | 0.6184 | 0.0017 | 1.856 | 1.262 | 2.729 |
| COPD | -0.1216 | 0.3466 | 0.886 | 0.687 | 1.141 |
| Hypertension | 0.1239 | 0.2142 | 1.132 | 0.931 | 1.376 |
| Diabetes mellitus | 0.0833 | 0.3824 | 1.087 | 0.902 | 1.310 |
| **BMI** |  |  |  |  |  |
| Unknown | -0.4645 | 0.0002 | 0.628 | 0.491 | 0.804 |
| 30≤ | -0.0545 | 0.6171 | 0.947 | 0.765 | 1.173 |
| **Admit source** |  |  |  |  |  |
| Home | 0.9060 | <0.0001 | 2.474 | 1.850 | 3.310 |
| Rehabilitation | 0.2904 | 0.2528 | 1.337 | 0.813 | 2.199 |
| Transfer from acute care hospital | 0.0544 | 0.8017 | 1.056 | 0.691 | 1.614 |
| **Oxygen saturation on presentation** |  |  |  |  |  |
| 80-89 | 0.6871 | <0.0001 | 1.988 | 1.511 | 2.616 |
| 90≤ | 0.9232 | <0.0001 | 2.517 | 1.962 | 3.230 |
| **Proning** |  |  |  |  |  |
| Yes | 1.1840 | <0.0001 | 3.267 | 2.667 | 4.003 |

Abbreviations: BMI, body mass index; COPD, chronic obstructive pulmonary disease

## DISCUSSION

This study represents a review of one of the largest cohorts of COVID-19 mortality that includes data documented in non-structured fields within the EHR. The experienced team of registered nurses was able to extract detailed information from the medical record that is typically not included in a structured dataset analysis. The demographics of the patients who died is similar to other published studies: age predominately over 69, male majority, payor mix [reflecting age and Medicare, along with a low number of self-pay (41; 1.6%)], and multiple comorbidities.^3-12^ This study provides a detailed clinical picture of the circumstances that precede the sudden deterioration in non-ICU patients reported by clinicians, but not fully examined in the literature. A striking feature of COVID-19 that has been reported is the rapid progression of of respiratory failure soon after the the onset of dyspnea and hypoxemia.^13^ The National Institutes for Health (NIH) has reported that hypoxemia is common in hospitalized patients with COVID-19 and that the criteria for hospital admission, intensive care unit (ICU) admission, and mechanical ventilation differ between countries.^14^ In some hospitals in the United States, >25% of hospitalized patients require ICU care, mostly due to acute respiratory failure.

Recommendations from the NIH are close monitoring for worsening respiratory status for adults with COVID-19 who are receiving supplemental oxygen. These recommendations are aligned with our findings in the non-ICU patient population.

Approximately 50% of deaths occurred at a non-ICU level of care, despite admission to the appropriate care setting with normal staffing. Our analysis of 1112 patients who experienced at least one RRT/CA at a non-ICU level of care revealed that 64.4% required an escalation in level of care. Of the 1112 RRT/CA patients, 664 (59.7%) presented to the hospital with oxygen saturation levels greater than or equal to 90%. In addition, 687 patients (61.8%) had no oxygen support. Of the RTT/CA patients 1031 (92.7%) were admitted to a non-ICU level of care with normal staffing levels which was appropriate based on their care needs. At presentation to the ED, the oxygen saturation levels for these patients were significantly higher than those for patients admitted to the ICU. Before the RRT/CA, the most recent oxygen saturation levels recorded for the non-ICU patients remained high at greater than or equal to 90% for 852 patients (76.7%). Oxygen saturations were documented within two hours of the RRT/CA in 454 (40.9%) patients in the RRT/CA cohort. When the RRT/CA was called, 479 patients (43.1%) had an oxygen saturation less than 80%, and 868 patients (78.1%) were on a non-rebreather/nonrebreather with nasal cannula. These data imply a sudden, unexpected deterioration in respiratory status requiring an RRT/CA call in a large number of non-ICU patients.

## LIMITATIONS

This study includes the following limitations. First, the study focuses on the demographic and clinical characteristics of in-hospital COVID-19 patients who died between March 13, 2020 and April 30, 2020; it does not provide a comparison group of similar patients who survived during the same time period. Second, data were obtained from the EHR and manually abstracted from medical records through retrospective review, but some routine documentation was less detailed due to the volume of patients being treated. Third, race was documented as other/unknown in 685 patients (26%); therefore, conclusions about race could not be drawn. Fourth, missing BMI data were included in the category of “Unknown” BMI. Finally, the study does not recognize a specific trigger that can distinguish which non-ICU patients in the cohort should be monitored.

## CONCLUSION

Patients admitted to a non-ICU level of care appear to suffer a rapid clinical deterioration, often with the hallmark of a sudden decrease in oxygen saturation. This finding suggests non-ICU patients could benefit from additional monitoring, such as continuous central oxygenation saturation. The availability of wireless patch monitoring should be considered, along with other methods, such as carbon dioxide and/or cardiac monitoring. Although this approach does not ensure reduced mortality, the number of RRT/CA calls infers this area warrants further study.

## Data Availability

The data that support the findings of this study are available on request from. The data are not publicly available due to restrictions as it could compromise the privacy of research participants.

## CONTRIBUTORS

MPJ had full access to all data in the study and takes responsibility for the integrity of the data and the accuracy of the data analysis. MPJ, SES, JSL, and KLN were responsible for the concept and design of the study. MPJ, SES, JSL, JJW, LS, MDG, and KLN were responsible for data acquisition, analysis, and interpretation. MPJ, SES, JSL, JJW, LS, and KLN were responsible for drafting the manuscript. MPJ, SES, JSL, JJW, LS, MDG, and KLN were responsible for critical revision of the manuscript for important intellectual content. JJW was responsible for the statistical analysis. MPJ, SES, JSL, JJW, LS, MDG, and KLN were responsible for administrative, technical, and material support. MPJ supervised the study.

## ACKNOWLEDGMENTS

We thank the Northwell Health Institute for Clinical Excellence & Quality/Safety: Mary Cama, RN, MSN; Patricia Meo, RN, BSN; Amy Logeman, RN, BSN, CPPS; Maureen McCarthy, RN, BSN; Josephine Fernandez-Kapilevich, RN, BSN; Theresa A. Droluk, RN, BSN; Jessica Martin, RN, BSN, RN-BC; Allison Carballo, RN, MBA; Jimmy Diaz, BS. We also thank the Northwell Health, Krasnoff Quality Management Institute: Alex Ma; Kahliik S. Burrell; Mark P. Tursi, MBA. We would like to acknowledge the contributions of the Northwell Health COVID-19 Research Consortium, including Crystal R. Herron, PhD, and Jennifer C. Johnson, MS, for editorial support. We acknowledge and honor all of our Northwell team members who consistently put themselves in harm’s way during the COVID-19 pandemic. We dedicate this article to them, as their vital contribution to knowledge about COVID-19 and sacrifices on the behalf of patients made it possible.

This work was supported by grants R24AG064191 from the National Institute on Aging of the National Institutes of Health and R01LM012836 from the National Library of Medicine of the National Institutes of Health.

